# Comparison of LSTM and Transformer Models for Activities of Daily Living Recognition using In-Home Ambient Sensors

**DOI:** 10.64898/2026.01.06.25342431

**Authors:** Nader Abdalnabi, Venkatanand Ram Addepalli, Shraboni Sarker, Andrew Kiselica, Erich Kummerfeld, Praveen Rao, Knoo Lee

## Abstract

This study evaluated Long Short-Term Memory (LSTM) and Transformer artificial intelligence (AI) models for recognizing Activities of Daily Living (ADLs) using data collected from a low-cost, non-invasive ambient in-home sensor system. Motion, temperature, luminance, and door-contact sensors were deployed in a two-participant home for 22 days, with ground truth established through volunteer logs and expert validation. Missing data were handled using Akima and linear interpolation. Models were trained using a 16/3-day train–validation split with the last three days reserved for testing to avoid temporal leakage. Performance assessments included participant-specific modeling, sequence-length variation, and hyperparameter tuning, using micro-accuracy and AUC-ROC as evaluation metrics. The Transformer consistently outperformed the LSTM, particularly for Participant 2 (AUC 0.91 vs. 0.87), and demonstrated superior adaptability to irregular time-series data. Findings underscore the feasibility of combining ambient sensing with AI for accurate ADL recognition and its potential to enable early detection of cognitive decline, reduce hospitalization risk, and alleviate caregiver burden.

## Introduction

The global aging population presents a significant challenge, with the number of U.S. adults aged 65 years and older projected to nearly double from 46.5 million in 2014 to 83.7 million by 2060^1^. The economic burden of aging-related illnesses, particularly Alzheimer’s disease and related dementias (ADRD), is projected to rise from $360 billion in 2024 to $1 trillion by 2050^2^. These disorders impose a substantial burden on patients, caregivers, family members and the broader healthcare system. Addressing the challenges posed by such demographic changes, including healthcare workforce shortages, requires innovative approaches^3^. Remote monitoring technology in the context of Alzheimer’s disease (AD) care offers new opportunities to alleviate caregiver burden and improve the quality of patient care^4^.

A Human Activity Recognition (HAR) system can monitor patient activities outside medical facilities, providing, offering non-invasive, passive in-home remote sensor system for continuous and accurate health tracking with minimal invasion of privacy^5, 6^. Such systems can also provide early warnings, enabling timely responses to health issues before they become more severe or costly^7^. Importantly, sensor systems extended the average independent housing stay by 1.7 years and saved $87,000 per person in projected Medicaid costs in one study^8^. While research on leveraging wearable sensors have shown progress on topics including classifying falls and other related risks, there remains gap disparities in adherence to true application of the technology, with older adults being lower affinity toward such wearable devices along with consent to data collection^9^. Furthermore, while using devices such as cameras for image recording or depth sensors for gait measurement which shown promising results ^10, 11^, the high cost of these devices limits their broader use^7^. Moreover, studies show that older adults are less likely to participate when a device or recording equipment is placed in their homes, since they see it as an invasion of privacy^11^. They also face challenges managing wearable sensors, even though these tools are widely used in remote health systems, which may lead to high attrition rates^12^. Therefore, there is a pressing need for cost-feasible, non-invasive solutions for assisting older adults that may overcome hesitation with implementation. In this study, we use cost-feasible, non-wearable sensors, offering a practical and unobtrusive solution for continuous in-home monitoring. Their low-dimensional design and minimal maintenance requirements make them well-suited for deployment in households of marginalized and aging populations.

We pair this sensor system with Machine learning (ML) and deep learning (DL) techniques for purposes of activity recognition. ML and DL are sub-branches of artificial intelligence (AI) have enabled breakthroughs in areas such as time series analysis and streaming daily activities recognition among many others. ML enables computers to learn patterns from data and improve performance on tasks without being programmed ^13^. DL employs multi-layered artificial neural networks to automatically learn hierarchical feature representations from large amounts of data ^14^. In recent years, researchers have employed ML and DL for classifying/recognizing Activities of Daily Livings (ADLs). <u>Al-gamdi et al., ^15^</u> showed that 1D Convolutional Neural Network (1d-CNN) outperformed Long-Short Term Memory (LSTM) models for recognizing ADLs in smart homes and achieved an F1 score of 0.90. Uday et al., ^16^ showed that Linear Support Vector Classifier (SVC) achieved better accuracy than Gated Recurrent Unit (96.7% vs. 92.60%) on smartphone data. <u>Sabbu et al., ^17^</u> used LSTM-based neural networks for recognizing human activities and achieved 100% accuracy albeit a small dataset. Onthoni et al., ^18^ used Gaussian Naive Bayes supervised learning for ADL recognition in a smart home environment achieving an accuracy of 85.30%. More recently, Augustinov et al., ^19^ showed that Transformer models can outperform LSTM models (73.36 % vs. 69.09 %) for recognizing ADLs from wearable sensor data. Cavalcante et al., ^20^ conducted a comparative study of DL techniques for recognizing ADLs using smartwatch data. The experimental results found an accuracy of 74.40 ± 20.40%, precision of 75.5 ± 19.70%, recall of 74.4 ± 20.40%, and F1-Score of 72.1 ± 20.90%. Other studies have optimized deep learning architectures and sensors data strategies to improve ADL recognition performance and scalability. For example, Wang et al., ^21^ proposed a deep learning model that combines convolutional layers and LSTM to enhance prediction accuracy, experiments conducted on two publicly available datasets and results indicated that concatenated representations of data generally outperform single representations. Fortino et al., ^22^ presented a novel technique for predicting daily living activities using a combination of Marked Temporal Point Processes and Neural Networks. It demonstrates significant improvements in prediction accuracy, outperforming existing methods by an average of 60% in F1-score across various datasets. Building on these findings, while the application of ML and DL techniques in sensor-based activity recognition has become widespread, most studies have primarily utilized high-dimensional or richly annotated datasets. Research focusing on low-dimensional sensor inputs (e.g., binary motion from infrared sensors, ambient temperature, and luminance) remains limited in predicting or classifying quality-of-life outcomes. With recent advances in AI, particularly transformer architectures, we hypothesize that transformer and LSTM models can achieve reliable activity classification for older adults using only low-dimensional sensor data, and we aim to compare their performance in this context.

## Materials and Method

### Data Source

Our study employed a set of non-wearable in-home sensors (i.e., ambient sensors that captures motion, temperature, and luminance; door sensor for entry/exit) to capture human activity, paired with Raspberry Pi 4G connected with Z-wave technology for data upload and connectivity (See Figure 1). This sensors system, developed by the University of Missouri^23^, has been deployed in multiple aging-in-place sites across the state. As shown in Figure 2, sensors were placed in key rooms (e.g., kitchen, living room, bathroom, bedroom) to capture motion, temperature, luminance, and entry/exit events. Devices communicated via a Z-Wave stick connected to a Raspberry Pi running the OpenHAB framework ^24^, with internet access provided through a Wi-Fi hotspot. The system was deployed in a home with two participants as shown in <u>Figure 2.</u> Sensor readings were transmitted to an external cloud server for secure storage and processing. These readings were made available through our custom dashboard for data usage. The collected sensor data were integrated and preprocessed to train and evaluate DL models for classifying ADLs. All study procedures were reviewed and approved by the University of Missouri Institutional Review Board (IRB #2101666).

**Figure 1.**
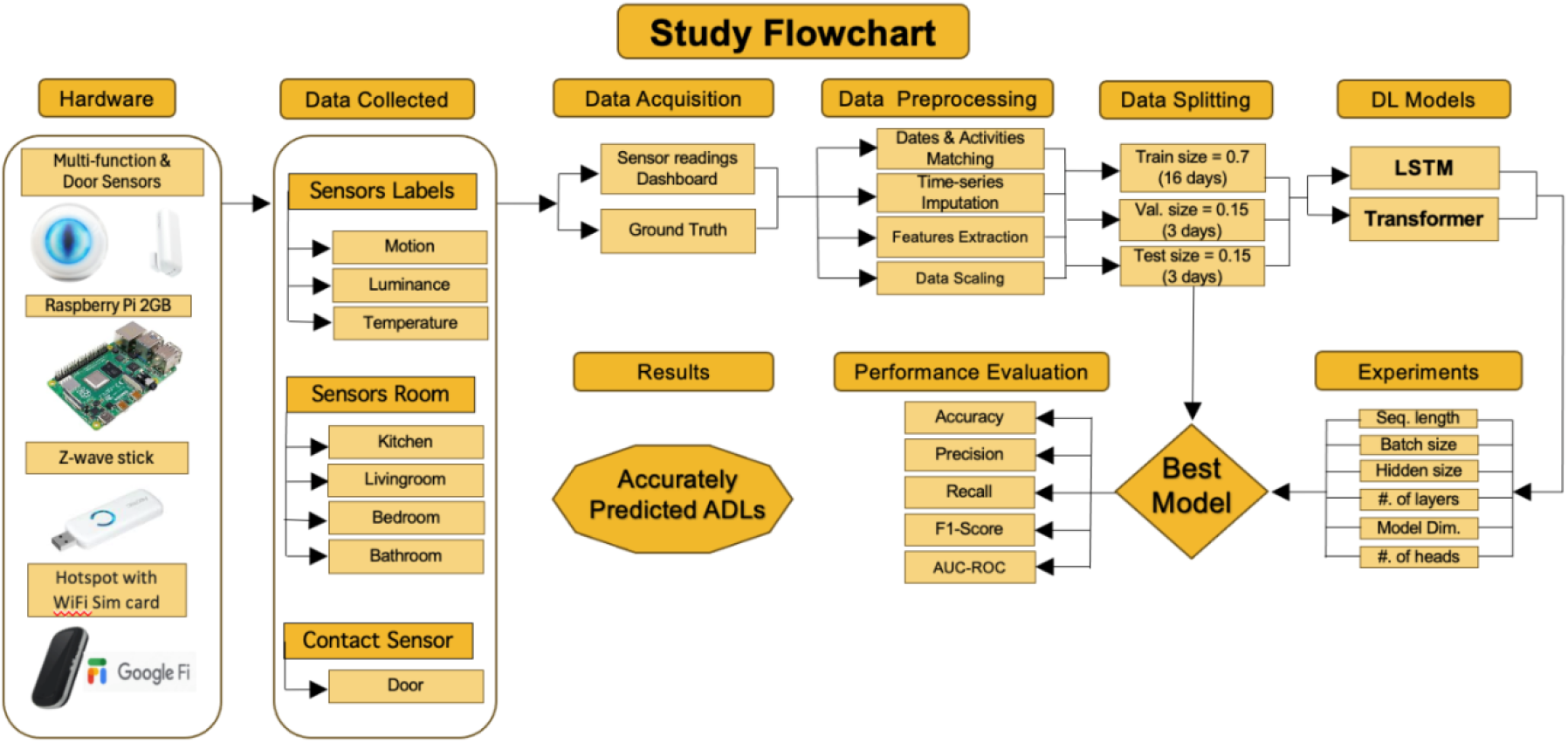
Study flowchart showing the ADL recognition pipeline. Non-wearable sensors collected motion, luminance, temperature, and door data across multiple rooms. After preprocessing and splitting into training, validation, and test sets, LSTM and Transformer models were trained with different hyperparameters. Model performance was evaluated using accuracy, precision, recall, F1-score, and AUC-ROC to select the best model. **Abbreviations DL:** Deep Learning **LSTM:** Long Short-Term Memory. **ADLs:** Activities of Daily Living. **AUC-ROC:** Area Under

**Figure 1.**
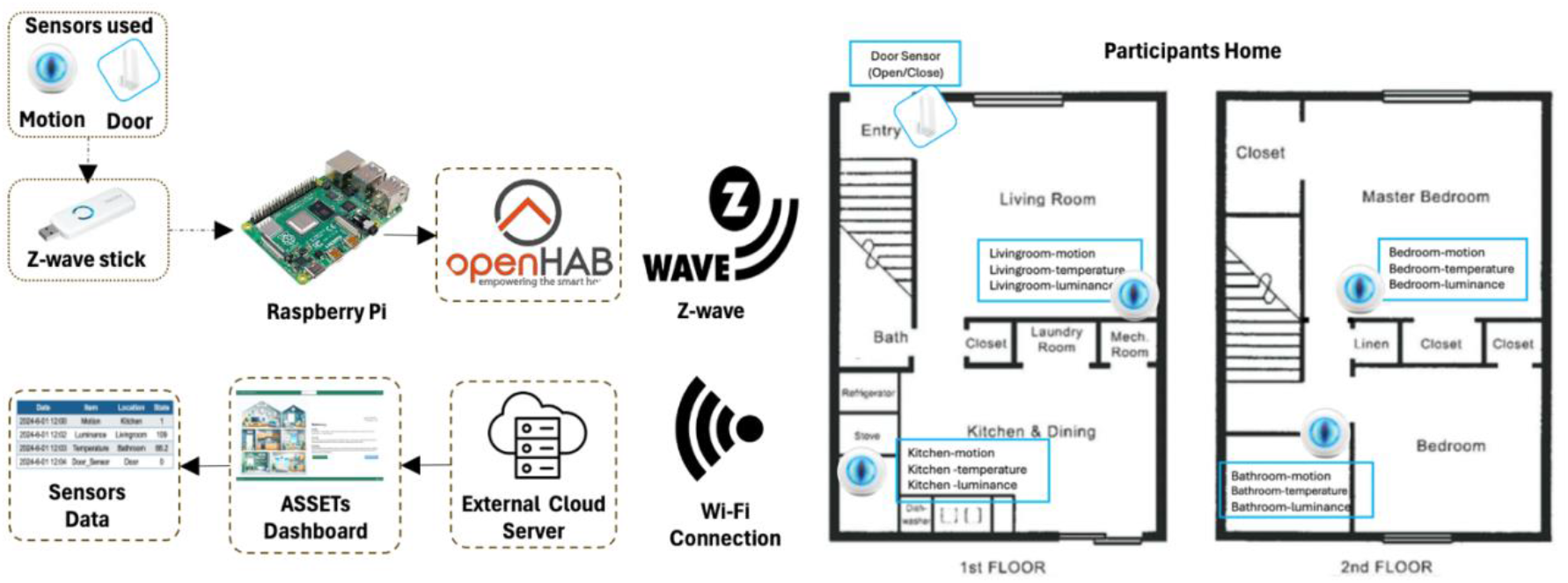
Overview of the smart home sensor deployment. Motion and door sensors connected via a Z-wave stick and Raspberry Pi transmitted data through the OpenHAB platform to a cloud server, accessible via the ASSETs dashboard. Sensors were placed across multiple rooms in the participant’s home, including the living room, kitchen, bedroom, and bathroom, to capture motion, temperature, and luminance signals for ADL recognition.

**Figure 2.**
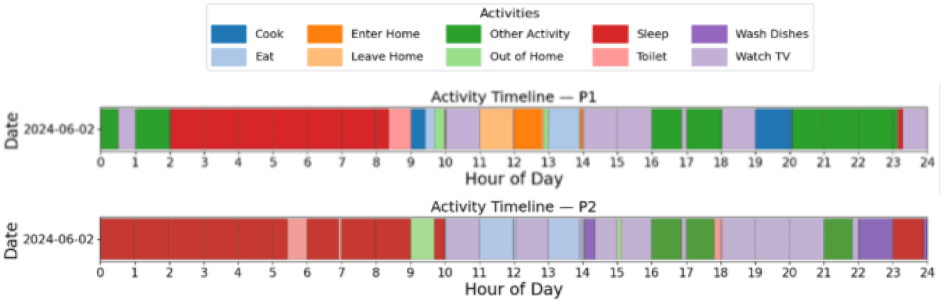
Participants’ activity timeline on June 2nd

### Input and Output Variables

Our input variables were based on the data collected by ambient sensor system included motion (binary), temperature (continuous), and illuminance (continuous). Motion events were recorded on an event-driven basis, while temperature and illuminance were reported according to the devices’ default operating modes either at fixed intervals (i.e., temperature – every 15 minutes; illuminance – every 60 minutes) or when a predefined threshold change was detected (i.e., 1 °C for temperature and ∼200 lux for illuminance).

The output variable was defined as Activities of Daily Living (ADLs) for model training, validation, and testing. Labels were assigned to core ADL tasks by mapping sensor-based events to activity categories, enabling supervised learning and performance evaluation. To align with clinically validated frameworks, ADLs were organized into three groups (i) Basic ADLs, following the Katz Index of Independence in Activities of Daily Living (i.e., bathing, dressing, toileting, transferring, continence, and eating)^25^; (ii) Instrumental ADLs (IADLs), based on the Lawton– Brody Scale (i.e., shopping, meal preparation, housekeeping, transportation, medication management, telephone use, and financial management)^26^; and (iii) Recreational/Other activities, designated as non-ADLs to avoid conflation with clinical measures. Table 1 lists the study main ADLs with associated activities and categories.

**Table 1.**
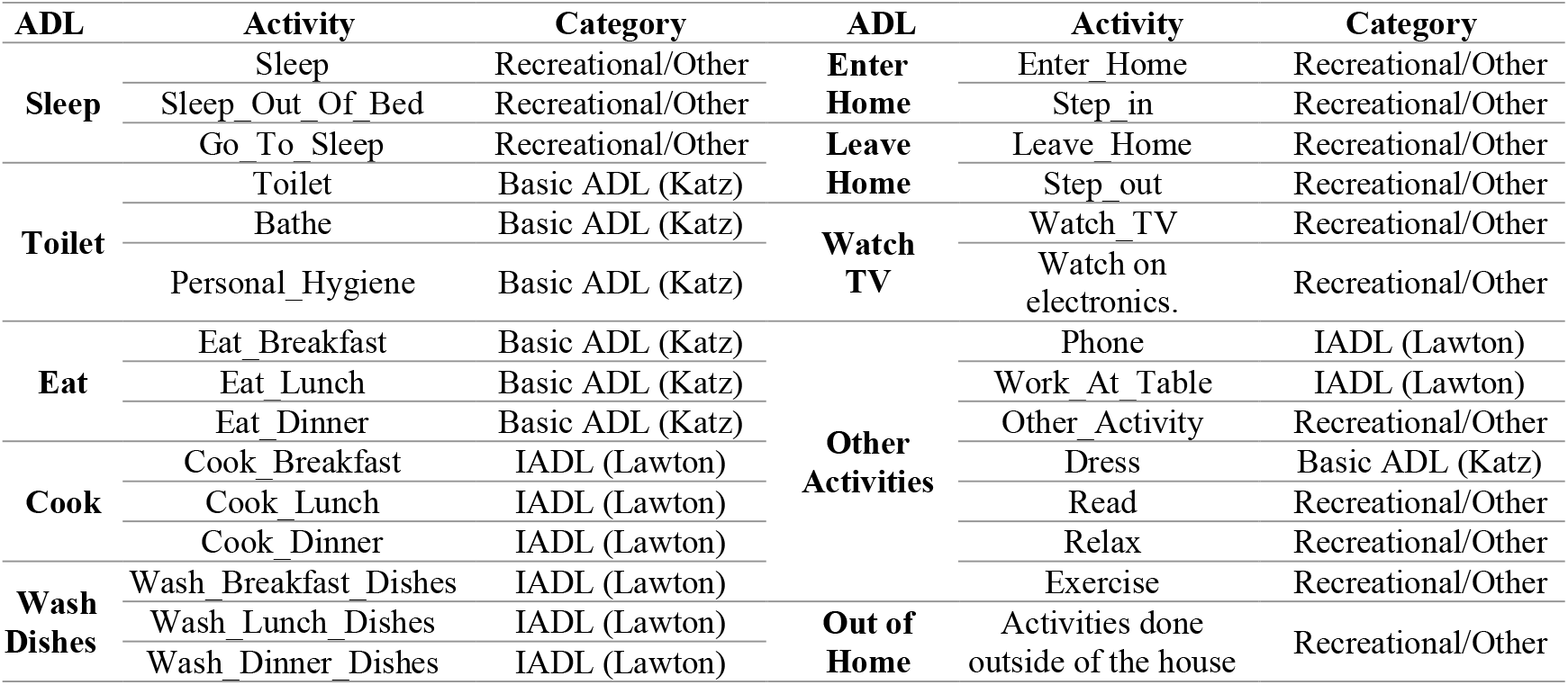
Study list of ADLs, activity and their categories.

ADLs were manually labeled by the two participants who recorded their ADLs on an hourly basis over a four-week period. These ADLs served as the output variable for model training, validation, and testing. Figure 3 illustrates a representative one-day for the hourly distribution of ADLs for both participants, highlighting routine behaviors and variability. Frequent activities such as Sleep, Watch TV, Other Activity and Out of Home activities dominate the dataset, while rarer activities (e.g., Cook, Eat, Wash Dishes) are underrepresented, underscoring the challenge of class imbalance in ADL recognition.

### Data Preprocessing

The preprocessing step aimed to produce a finalized time series sensors dataset with accurately aligned ADL labels. Data handling was performed in Python using pandas for timestamp standardization. To align logs with sensor events, hourly self-reported activities were fuzzy matched to minute-level sensor data within a defined window. Rows with multiple activities were resolved by retaining the entry most relevant to the corresponding room and sensor, based on an expert-derived mapping. Because Transformer models require ordered temporal input, gaps and irregularities in the sensor streams were interpolated to maintain a consistent 1-minute interval. For temperature and luminance, both linear and Akima interpolations were tested; as shown in Figure 4 (B) below, Akima produced smoother values while preserving the overall trends and directional patterns of the original data than linear interpolation in Figure 4 (A), Akima also improved model performance as it will be shown later in the Results section. For motion or door data (binary data), forward-filling, a technique to handle missing categorical sequential data by replacing a gap with the last known preceding value, was applied assuming the last state persisted until a new event occurred.

**Figure 4.**
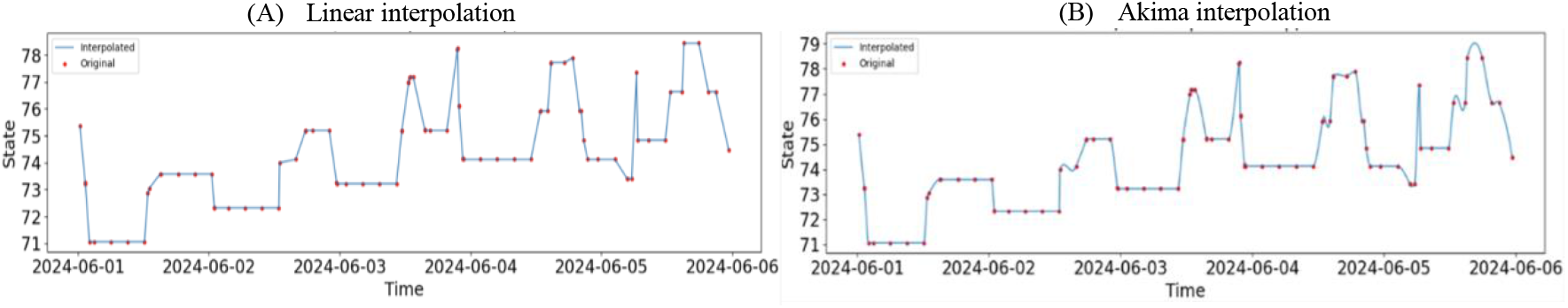
Comparison of interpolation methods for temperature sensors time-series data (A) Linear interpolation (left) versus (B) Akima interpolation (right). Akima produced smoother values yielding better model performance.

Feature engineering ensured a structured representation for model input. Continuous (temperature & luminance) and binary (motion & door) states were separated into distinct features, and room identifiers (i.e., kitchen, living room, bedroom, bathroom and door) were extracted. Table 2 shows a sample of the final preprocessed dataset used to train the DL models.

**Table 2.** Sample of sensor readings with corresponding ADL labels for the two participants.

| Date | Door | Light Sensors |  |  |  | Motion Sensors |  |  |  | Temperature Sensors |  |  |  | Participants |  |
| --- | --- | --- | --- | --- | --- | --- | --- | --- | --- | --- | --- | --- | --- | --- | --- |
|  | Door | Bath | Bed | Kitchen | Living | Bath | Bed | Kitchen | Living | Bath | Bed | Kitchen | Living | P1 | P2 |
| 2024-06-01 17:58:00 | 0.00 | 3.84 | 538.60 | 17.59 | 367.80 | 1.00 | 0.00 | 0.00 | 0.00 | 73.58 | 71.12 | 74.30 | 70.49 | Toilet | Watch_TV |
| 2024-06-01 17:59:00 | 0.00 | 3.88 | 539.86 | 17.46 | 369.00 | 1.00 | 0.00 | 0.00 | 0.00 | 73.58 | 71.16 | 74.30 | 70.50 | Toilet | Watch_TV |
| 2024-06-01 18:00:00 | 0.00 | 3.91 | 540.93 | 17.33 | 369.91 | 1.00 | 0.00 | 0.00 | 0.00 | 73.58 | 71.21 | 74.30 | 70.51 | Toilet | Watch_TV |
| 2024-06-01 18:01:00 | 0.00 | 3.95 | 541.78 | 17.20 | 370.51 | 0.00 | 0.00 | 1.00 | 1.00 | 73.58 | 71.25 | 74.30 | 70.52 | Cook | Eat |
| 2024-06-01 18:02:00 | 0.00 | 3.97 | 542.41 | 17.07 | 370.86 | 0.00 | 0.00 | 1.00 | 1.00 | 73.58 | 71.29 | 74.30 | 70.53 | Cook | Eat |
| 2024-06-01 18:03:00 | 0.00 | 4.00 | 542.80 | 16.93 | 370.96 | 0.00 | 0.00 | 0.00 | 0.00 | 73.58 | 71.33 | 74.30 | 70.53 | Cook | Eat |
| 2024-06-01 18:04:00 | 0.00 | 4.02 | 542.95 | 16.79 | 370.87 | 0.00 | 0.00 | 0.00 | 1.00 | 73.58 | 71.38 | 74.30 | 70.54 | Eat | Eat |
| 2024-06-01 18:05:00 | 0.00 | 4.05 | 542.83 | 16.65 | 370.60 | 0.00 | 0.00 | 0.00 | 1.00 | 73.58 | 71.43 | 74.30 | 70.55 | Eat | Eat |
| 2024-06-01 18:06:00 | 0.00 | 4.06 | 542.46 | 16.50 | 370.19 | 0.00 | 0.00 | 1.00 | 1.00 | 73.58 | 71.47 | 74.30 | 70.56 | Eat | Wash_Dish |

### Sampling and Model Training

Our dataset contained sensor readings for 22 days. For model training, we used data for 19 days with an 85/15 train– validation split i.e. 16 days for training set, 3 days for validations set. Data of the last three days (≈15% of the 22-day dataset) was used as the hold out test set. Three experiment sets were conducted: a) In the first experiment, we trained the LSTM and Transformer models for predicting a participant’s ADL with and without the other participant’s ADL included as feature during training. b) In the second experiment, we varied the sequence length (i.e., number of consecutive sensor readings/rows in the input) for the DL models. We generated the input sequences in a sliding window manner by restricting them to be within single days to avoid cross-day leakage. The sequence length seq_len ∈{5, 10, 20, 25, 30}. The target was the ADL label of the last record/reading in the sequence. c) In the third experiment, we changed the model hyperparameters. We tested the following settings: batch size b∈{16, 32}; LSTM parameters hidden size ∈{16, 32}; number of layers ∈{1, 2}; Transformer parameters encoder-only architecture with hidden size ∈{16, 32}; number of layers ∈{1, 2}; and attention heads ∈{4, 8}. Fixed parameters included learning rate = 1e-3, dropout = 0.2, and 100 epochs with early stopping (patience = 10).

Model performance was assessed using micro-averaged at 0.5 threshold accuracy, precision, recall, and F1-score. Micro-averaging was chosen over macro-averaging to address class imbalance in our dataset, as it aggregates contributions from all classes based on their frequency, therefore, ensure minor classes (i.e. eat, cook) representation. In addition, the Area Under the Curve of the Receiver Operating Characteristic (AUC-ROC) score was computed to measure a models’ ability to separate ADL classes. AUC is particularly valuable in multi-class ADL prediction, because it is less sensitive to class imbalance and provides a robust indicator of the DL models’ discriminative capacity across both frequent and infrequent activities ^27^.

Experiments were run on a high-performance computing machine (Linux OS, NVIDIA GeForce RTX 4090 GPU) using Python 3.12.7 with PyTorch 2.6.0+cu124 and CUDA 12.4. Supporting packages included NumPy 1.26.4, Pandas 2.2.2, Matplotlib 3.9.2, and Scikit-Learn 1.5.1 for preprocessing, visualization, and evaluation.

## Results

### Data Overview

The raw sensors dataset contained 16,384 rows, which increased to 31,536 rows after Akima interpolation as shown in Table 3. Activity distributions differed between the two participants, reflecting variation in ADL patterns. For Participant 1, the most frequent activity was Out of Home (56.4%), followed by Sleep (19.9%) and Other Activity (12.1%). Less common activities included Watch TV (4.6%), Leave Home (1.9%), Enter Home (1.8%), Eat (1.2%), Cook (1.1%), and Toilet (0.9%), with no Wash Dishes activity. For Participant 2, Sleep was the most frequent activity (37.7%), followed by Out of Home (31.3%), Other Activity (11.6%), and Watch TV (7.9%). Lower-frequency activities included Eat (3.1%), Enter Home (2.6%), Toilet (2.2%), Leave Home (2.1%), Cook (0.8%), and Wash Dishes (0.6%). Overall, P1 spent a greater proportion of time outside the home, while Participant 2 spent more time sleeping and engaging in in-home activities. The strong class imbalance across both participants highlights the need for evaluation metrics such as micro-averaged accuracy and AUC, which better capture overall model performance in the presence of uneven activity distributions. The activity distributions differed between the two participants due the fact that the data were collected in a regular home environment without over-controlled settings, so that the activity patterns more closely resemble real-life behaviors.

**Table 3.** Study data target classes distribution.

| Activity | Participant 1 |  | Participant 2 |  |
| --- | --- | --- | --- | --- |
|  | Count | % | Count | % |
| Out of Home | 17,797 | 56.43 | 9,878 | 31.32 |
| Sleep | 6,264 | 19.86 | 11,889 | 37.70 |
| Other Activity | 3,827 | 12.14 | 3,664 | 11.62 |
| Watch TV | 1,453 | 4.61 | 2,500 | 7.93 |
| Leave Home | 600 | 1.90 | 667 | 2.12 |
| Enter Home | 570 | 1.81 | 815 | 2.59 |
| Eat | 378 | 1.20 | 967 | 3.07 |
| Cook | 356 | 1.13 | 264 | 0.84 |
| Toilet | 291 | 0.92 | 699 | 2.22 |
| Wash Dishes | 0 | 0.00 | 193 | 0.61 |
| **Total** | **31,536** | **100** | **31,536** | **100** |

#### LSTM Model

Table 4 (top) summarizes the best five LSTM configurations for Participant 1 (P1) and Participant 2 (P2), ranked by AUC rounded to two decimal points. For P1, the top three models (AUC = 0.95) achieved accuracies between 0.71 and 0.74. The highest-performing configuration used a sequence length of 20, batch size of 32, one hidden layer, and hidden size of 32, when P2’s data were included (accuracy = 0.73). Comparable AUC values (0.95) were obtained without P2’s data using sequence lengths of 10 or 20 and hidden sizes of 16–32, yielding accuracies of 0.71–0.74. The remaining two models achieved AUC = 0.94 with accuracies of 0.71–0.74 using similar parameter settings. For P2, the best configurations (AUC = 0.87) achieved accuracies of 0.56 under setups with sequence lengths of 10, batch size 32, and hidden size 32, either with one or two hidden layers. Other settings, including sequence lengths of 10 or 20 and hidden sizes of 16–32, maintained AUC values between 0.85 and 0.86, with accuracies ranging from 0.43 to 0.52.

**Table 4.**
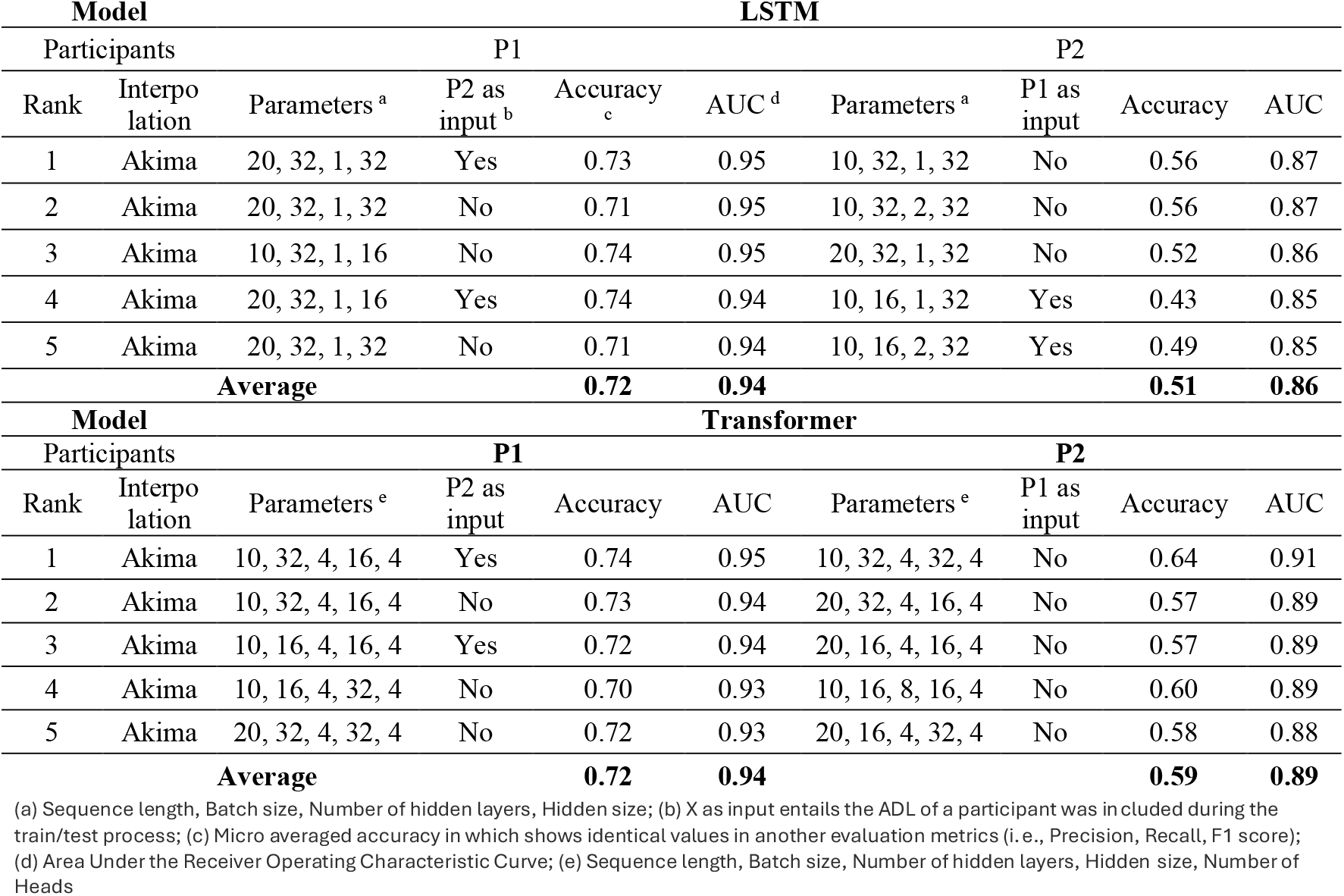
Comparison of LSTM and Transformer model performances for Activity of Daily Living (ADL) recognition across Participants 1 (P1) and 2 (P2). Each model was evaluated with and without cross-participant input inclusion (“P2 as input” for P1 and “P1 as input” for P2). Accuracy at a threshold of 0.5. Results are ranked by AUC. Average metrics are shown at the bottom of each model section.

#### Transformer Model

Table 4 (bottom) presents the best five Transformer configurations for P1 and P2, also ranked by AUC rounded to two decimals. For P1, the top model achieved an AUC of 0.95 and an accuracy of 0.74 using sequence length 10, batch size 32, four layers, hidden size 16, and four attention heads, when P2’s data were included. Two other high-ranking models achieved AUC = 0.94 with accuracies between 0.72 and 0.73 under similar configurations with sequence lengths of 10 and batch sizes of 16–32. The remaining models produced AUC = 0.93 and accuracies between 0.70 and 0.72 using sequence lengths of 10–20 and hidden sizes of 32. For P2, the top-ranked configuration (AUC = 0.91) reached an accuracy of 0.64 with sequence length 10, batch size 32, four layers, hidden size 32, and four heads, without including P1’s data. Other configurations achieved AUCs of 0.88–0.89 and accuracies between 0.57 and 0.60, demonstrating consistently steady classification across varying sequence lengths and hidden sizes.

## Discussion

This study explored the feasibility of using data collected by ambient non-wearable, low-cost sensor system for motion, luminance, temperature, and door-contact sensors, and deep learning models (i.e., LSTM and Transformer) to recognize ADLs in adults.

For P1, both the LSTM and Transformer models achieved similar levels of accuracy at a threshold of 0.5, but exhibited distinct sensitivities to their parameter configurations. The LSTM achieved its top performance (accuracy = 0.73, AUC = 0.95) using longer sequences (SeqLen = 20), a single hidden layer and larger hidden sizes (32). This indicates that extended temporal windows (SeqLen) with richer internal representation (hidden size) helped the model capture subtle transitions in activity sequences. The Transformer, in contrast, achieved its best accuracy (accuracy = 0.74, AUC = 0.95) with shorter sequences (SeqLen = 10), larger batch size = 32, smaller hidden size = 16, and four layers with four attention heads. This highlights the Transformer’s efficiency in learning from shorter temporal windows and larger batch sizes due to advances structure which regularized attention across time steps. While both models benefited from including cross-participant data (P2), the Transformer’s higher AUC across configurations reflects superior discriminative ability thanks to Transformer’s attention mechanism, achieving high performance without relying on long sequences or small batches.

For Participant 2 (P2), the performance gap between the LSTM and Transformer models was noticeable. The Transformer achieved a higher accuracy of 0.64 and an AUC of 0.91, compared to the LSTM’s best accuracy of 0.56 and AUC of 0.87, demonstrating stronger class separability and overall discriminative capability. The LSTM showed improved performance, models 4 & 5, when P1’s data were included, suggesting that it relied on the additional contextual variability to learn temporal dependencies across participants. In contrast, the Transformer achieved its best performance without P1’s data, highlighting its ability to generalize effectively and extract robust temporal representations from P2’s data alone. The LSTM tended to perform better with longer input sequences (SeqLen = 20), smaller batch sizes (16), and hidden sizes around 32, again, reflecting its dependence on extended temporal context and gradual sequence learning. The Transformer, however, performed best with shorter sequences (SeqLen = 10), larger batch sizes (32), and four layers, indicating that its attention mechanism can selectively focus on the most informative time steps without requiring long input windows. Across all tested configurations, Transformer AUC values (0.88–0.91) remained consistently higher than those of the LSTM (0.85–0.87), underscoring the robustness of self-attention in handling class imbalance and overlapping activity patterns. Overall, while the LSTM benefited from longer sequences and cross-participant input, the Transformer demonstrated superior adaptability, achieving higher accuracy and AUC with fewer temporal dependencies.

Clinically, our results highlight the potential of ADL recognition for detecting rare but meaningful changes such as mobility decline or shifts in self-care routines ^28^. These subtle deviations often serve as early indicators of Alzheimer’s disease and related dementias (ADRD), where disruptions in daily activities can precede more severe memory loss, behavioral changes, and functional impairment ^29 30^. Early detection through continuous ADL monitoring enables timely interventions that may slow deterioration and support independent living. Additionally, studies have shown that home-based monitoring reduces hospital readmissions, emergency department visits, and overall hospital days, with one trial reporting hospitalizations dropping from 0.45 to 0.19 at 3 months and from 0.55 to 0.23 at 6 months, alongside significant reductions in ED visits ^31 32^. These findings altogether underscore that ADL recognition, particularly the Transformers with superior class separability, has the potential to facilitate early detection of cognitive decline, reduce preventable hospitalizations, and ease healthcare providers’ burden.

Moreover, ADL recognition systems may lessen the stress and burden on caregivers and family members by reducing direct impact, guilt, and frustration/embarrassment ^33^. Our results highlight participant-specific variability i.e. ADL recognition for P1 performed best with shorter sequences and smaller hidden sizes, while ADL recognition in P2 benefited from longer sequences and larger hidden sizes. This variability underscores the need for AI models to be tuned to individuals, linking directly to personalized monitoring. A well-tuned and personalized model reduces false alarms and builds caregiver trust, thereby lowering stress and improving system usability. Such options may also create opportunities for tailored personalized interventions e.g. rehabilitation, nutrition, or exercise programs ^34 35^.

Our study demonstrates that ambient nonwearable, low-cost sensors combined with deep learning models can provide effective ADL recognition without relying on wearables devices, which have concerns regarding data quality, balanced estimations, health equity, and fairness ^36^. This design removes the burden of remembering to wear or recharge devices. This change is especially important for older adults with cognitive decline, while also ensuring affordability and scalability for underserved populations. Our study situates itself within the broader trends of applying AI in remote patient monitoring (RPM) and smart healthcare ecosystems. It complements advances in federated learning, which can safeguard privacy while building ADL models across distributed homes ^37 38^, and reinforcement learning, which may enable adaptive, personalized interventions that evolve with patient needs ^39^.

## Future Work

Future research should explore adaptive hyperparameter tuning, as our results show that some participants benefit from shorter sequences and smaller hidden sizes, while others perform better with longer sequences and larger hidden sizes. Building adaptive learning frameworks that automatically optimize these parameters per user could enhance personalization and system reliability. Moreover, since Transformers demonstrated superior class separability, they can be extended to detect rare but clinically significant events such as falls or early indicators of cognitive decline. In addition, incorporating federated and reinforcement learning offer a promising path to train models across diverse participants while preserving privacy, especially given the cross-participant effects observed in our study. These directions can strengthen ADL recognition systems, making them more personalized, privacy-preserving, and clinically impactful.

## Limitations

Despite promising results and a practical sensor system, our study faced several limitations. First, the dataset was collected from a limited number of homes, which may restrict generalizability. However, small-scale deployments are common in early-stage ADL recognition research, as they allow for controlled experimentation, feasibility testing, and iterative improvement before scaling to larger populations. Second, door sensor data represented only about 2% of the dataset, raising concerns about statistical significance. However, including this data was important for capturing transitional activities (e.g., entering and leaving home). Although these activities are infrequent, they are clinically relevant for assessing independence and mobility. Third, factors such as sensor noise, coverage limits, and variability in sensor placement may have influenced the readings. These issues are inherent to real-world smart home environments and reflect the natural variability that participants would experience in their daily lives. Finally, contextual factors such as daily schedules, environmental changes, or weather were not explicitly modeled. While this may have influenced luminance and temperature readings, our goal was to first establish baseline feasibility using unobtrusive, low-cost sensors before incorporating more complex contextual information.

## Conclusion

In this study, we demonstrated that ambient, non-wearable, cost-feasible sensors combined with DL models can effectively support ADL recognition in real-world home environments. LSTM models showed consistent accuracy, while Transformer models provided stronger class separability with similar accuracy, which is critical for identifying rare but clinically meaningful activities. Participant-specific variability in optimal sequence length and hidden size highlights the importance of individualized parameter tuning to improve trust and reduce false alarms. These findings suggest that ADL recognition systems can contribute to earlier detection of cognitive and functional decline, reduced hospital utilization, and relief of caregiver burden. Future work should focus on adaptive learning frameworks, rare-event detection, and federated approaches to ensure scalability, privacy, and equity in deployment.

## Data Availability

All data produced in the present study are available upon reasonable request to the authors

## Acknowledgments

The authors also like to acknowledge the feedback from Dr. Blain Reeder.

